# The fate of SARS-CoV-2 in WWTPs points out the sludge line as a suitable spot for monitoring

**DOI:** 10.1101/2020.05.25.20112706

**Authors:** Sabela Balboa, Miguel Mauricio-Iglesias, Santiago Rodriguez, Lucía Martínez-Lamas, Francisco J. Vasallo, Benito Regueiro, Juan M. Lema

**Author notes:** Equal contribution (Sabela Balboa, Miguel Mauricio-Iglesias).

## Abstract

SARS-CoV-2 genetic material is detectable in the faeces of a considerable part of COVID-19 cases and hence, in the urban wastewater. This fact was confirmed early during the spread of the COVID-19 pandemic and prompted several studies that proposed monitoring its incidence by wastewater. This paper studies the fate of SARS-CoV-2 genetic material in wastewater treatment plants using RT-qPCR with a two-fold goal: i) to check the safety of the water effluent and also of the sludge produced and ii) based on the understanding of the virus particles fate, to identify the most suitable spots for detecting the incidence of COVID-19 and monitor its evolution. On the grounds of the affinity of enveloped virus towards biosolids, we hypothesized that the sludge line acts as a concentrator of SARS-CoV-2 genetic material. Sampling several spots in primary, secondary and sludge treatment at the Ourense (Spain) WWTP showed that, in effect, most of SARS-CoV-2 particles cannot be detected in the water effluent as they are retained by the sludge line. We identified the sludge thickener as a suitable spot for detecting SARS-CoV-2 particles thanks to its higher solids concentration (more virus particles) and longer residence time (less sensitive to dilution caused by precipitation). Although more studies will be needed for confirmation, these results contribute to clarify the role of WWTPs in COVID-19 mitigation.

## Introduction

Urban wastewater can be a vector for the spread of viral diseases, especially viruses that are transmitted through the faecal-oral route. There are numerous descriptions of virus detection in wastewater plants, including Norovirus, Sapovirus, Hepatitis A virus, Adenovirus, Poliovirus or Enterovirus among others (Ehlers et al., 2005; Sassi et al., 2018; Symonds et al., 2016; Taboada-Santos et al., 2020). Current knowledge regarding the behaviour of SARS-CoV-2 in wastewater is very limited, although its RNA has been detected in faeces of symptomatic individuals (Holshue et al., 2020; Woelfel et al., 2020) and in urban wastewater in different countries (Medema et al., 2020; Randazzo et al., 2020a, 2020b; Rosa et al., 2020; Wu et al., 2020; Wurtzer et al., 2020). Although it is assumed that the enveloped viruses are not excreted in high concentrations and that their survival in water is limited, there is little experimental evidence to confirm these hypotheses in wastewater; in fact, transmission of these viruses through wastewater was identified as responsible for an outbreak of severe acute respiratory syndrome (SARS) in Hong Kong in 2003 (Yu et al., 2004).

Urban wastewater constitutes a complex matrix which includes suspended solid materials, colloidal and dissolved biodegradable organic matter, nutrients, pathogens, etc. In wastewater treatment plants (WWTP) most of solids are separated from the water to the so-called sludge line. The vast majority of WWTPs have a first stage of homogenization and separation of solid (primary settler) and a secondary settler where the activated sludge is separated from the clarified water. Finally, the two types of sludge (primary and secondary) are concentrated in the thickener from where they are sent to the sludge treatment unit.

Pollutants of hydrophobic nature are mostly retained in primary or secondary sludge (Prado et al., 2014), a phenomenon described already many decades ago (Wellings et al., 1976). It is known that enveloped viruses, due to the presence of a lipid bilayer surrounding the protein capsid, have a different affinity to non-enveloped viruses, with a greater tendency to adsorb to solid and/or colloidal particles (Ye et al., 2016). This was experimentally proved to occur for two enveloped viruses: Murine coronavirus MHV (murine hepatitis virus) and *Pseudomonas* phage ϕ6 (Ye et al. 2016). Therefore, most probably, SARS-CoV-2 and particles thereof are indeed hydrophobic and, accordingly, they would be associated to the solids and/or colloidal material. Yet, most of the current literature concerning SARS-CoV-2 or its genetic material in wastewater deals with their presence in the water phase and very little attention has been paid to their fate in the sludge line.

Another aspect of concern for water boards and utilities is the potential transmission of SARS-CoV-2 in WWTP and their effluents. Actually, what is known so far about the transmission of SARS-CoV-2 is not particularly worrisome, given that WWTP operation is already intended to avoid the transmission of potential pathogens present in wastewater. Being sludge and the water effluents the main outflows from a WWTP, it is important to ascertain whether SARS-CoV-2 can be detected in these streams, even keeping in mind that the presence of SARS-CoV-2 genetic material does not imply that it is in an infective state.

Hence, this manuscript pays special attention to the sludge in WWTPs. The sludge treatment is very heterogeneous in WWTPs as large plants often feature anaerobic digestion treatment, usually at moderate temperatures (35-40°C) and long residence times (10-20 days), which would help to inactivate the possible viral load. In contrast, in smaller plants sludge can receive a mere heat drying treatment before being shipped to an authorized manager, or even just centrifugation to reduce its water content.

The goal of this manuscript is two-fold: first, to shed some light on the fate of SARS-CoV-2 in WWTPs by examining the detection of its genetic material along the water and sludge lines and second to check whether the hydrophobic nature of SARS-CoV-2 make the sludge line, and particularly the thickened sludge, as a suitable spot for monitoring its incidence in the WWTP catchment area.

## Materials and methods

### Wastewater and sludge samples

Wastewater and sludge samples were taken from Ourense WWTP in north-western Spain (characteristics and sampling points in Table 1). The 250 ml samples were taken twice a week from April 6 to April 21 2020, kept at 4°C before being sent in less than 24 h to the Universidade de Santiago de Compostela (USC) facilities to be concentrated.

**Table 1.** Wastewater treatment plant characteristics and sampling points for wastewater and sludge

| **Ourense WWTP Characteristics** |  |
| --- | --- |
| Nominal size | 350 000 p. equivalent |
| Pretreatment | Grit and sand separator, oil and grease removal |
| Wastewater treatment | Primary settler<br>Biological SBR for COD and N removal<br>Chemical removal of P<br>Microfiltration of secondary effluent |
| Sludge treatment | Gravity thickening and centrifuge, thermal hydrolysis, anaerobic digestion |
| Sampling points water line | Wastewater after grit removal; outflow primary settler; outflow secondary treatment |
| Sampling points sludge line | Primary sludge, secondary, thickened mixed sludge, digested sludge |

For the water line, 24-h composite samples were taken and characterised in terms of pH, conductivity, total and volatile suspended solids (Standard Methods 2540), chemical oxygen demand (spectrophotometry, Standard Methods 5220-D), ammonium (spectrophotometry, Standard Methods 4500-F), nitrate (kit equivalent to DIN 38405-9), total nitrogen (kit equivalent to ISO 11905-1) and phosphorus (spectrophotometry, 4500-P).

**Figure 1.**
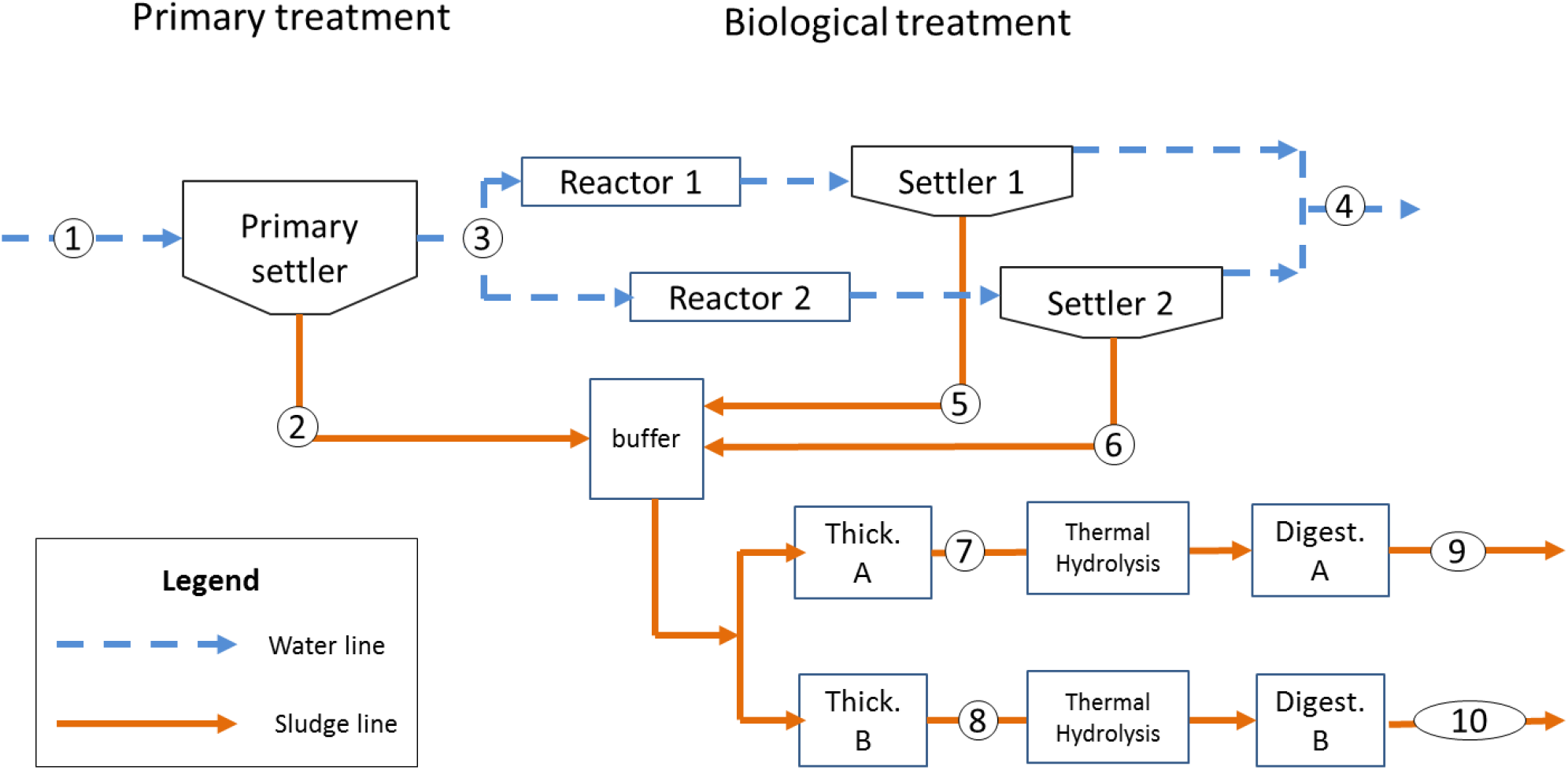
Simplified description of Ourense WWTP layout and sampling points in the water and sludge line

### Sample processing

Water samples were concentrated by ultrafiltration. Briefly, 100 ml were gently centrifuged to remove large particles at 4600 xg during 30 min. Supernatants were concentrated by filtration using Amicon 15 ml 10 K centrifugal devices and buffer was exchanged to phosphate buffer saline (PBS) pH 7.4.

Sludge samples were concentrated by precipitation with polyethylene glycol (PEG) according to Hjelmso et al. (2017). Then, 1:8 (v/v) of Glycin buffer (0.05 M glycine, 3% beef extract) was added to 50 ml of sludge, incubated for 2h at 4°C to detach viruses bound to organic material. Samples were then centrifuged at 8 000 xg during 30 min and filtered through a 0.45 μm polyethersulfone (PES) membrane to remove eurkaryotic and prokaryotic cells. Then, viruses were precipitated by adding 1:5 (v/v) of PEG 8000 (80g/L) and NaCl (17.5 g/L) during an overnight shacking (150 rpm) at 4°C and a centrifugation at 13 000 xg during 90 min. Samples were then resuspended in PBS buffer pH 7.4 and stored at -80°C for further analysis. Concentration control was performed with bacteriophage MS2, by inoculating each sample with 250 μl the virus (5.5 × 10^6^ viral particles/ml).

### RNA extraction and RT-qPCR detection

RNA extraction and RT-qPCR was carried out at the department of Microbiology of *Complexo Hospitalario Universitario de Vigo*. Nucleic acid extraction from both water and sludge concentrated samples was performed using MicrolabStarlet IVD using the STARMag 96 × 4 Universal Cartridge Kit (Seegene, Seoul, South Korea) according to manufacturer specifications.

Viral RNA was detected and quantified by a one-step multiplex RT-qPCR Allplex system™ 2019-nCoV (Seegene, Seoul, South Korea). The assay is designed to detect RNA-dependent polymerase (RdRP) and nucleocapsid (N) genes specific to SARS-CoV-2, and a region conserved in the E gene of the structural protein envelope for the detection of pan-Sarbecoviruses including SARS-CoV-2. The test uses internal RNA control for sample preparation and control of the PCR amplification process. For the RT-PCR, the CFX96 system was used ™ (Bio-Rad Laboratories, Hercules, CA, USA). The analysis of the results was performed using the specific Seegene viewer software 2019-nCoV. In parallel, a SARS-CoV-2 EDX standard (Bio-Rad, Hercules, CA) containing synthetic RNA transcripts of five SARS-CoV-2 genetic targets (genes E, N, ORF1ab, RdRp and S) of known concentration was used to establish a linear regression curve and obtain the concentration in copies/ml.

## Results and discussion

### Wastewater Physicochemical Characterization

The characterisation of the sample in terms of chemical oxygen demand (COD), suspended solids and, for water samples, total nitrogen is shown in Table 2. It can be seen how the influent has a very variable composition, which is most probably caused by precipitation events.

**Table 2.** Physicochemical characterisation of samples (full characterisation in supplementary materials). NA = Non-available sample

| Sampling spot |  | 6-Apr | 7-Apr | 14-Apr | 16-Apr | 21-Apr |
| --- | --- | --- | --- | --- | --- | --- |
| Influent (1) | COD (mgO/L) | 1211 | NA | 613 | 535 | 387 |
|  | TSS (mg/L) | 990 | 100 | 280 | 350 | 230 |
|  | NT (mgN/L) | 56 | 21 | 52 | 38 | 37 |
| Outflow Primary (3) | COD (mgO/L) | 101 | 53 | 212 | 160 | 59 |
|  | TSS (mg/L) | 51 | 75 | 128 | 107 | 33 |
|  | NT (mgN/L) | 35 | 26 | 41 | 38 | 30 |
| Treated Effluent(4) | COD (mgO/L) | 17 | 13 | 23 | 19 | 13 |
|  | TSS (mg/L) | 2.5 | 6.0 | 8.6 | 3.0 | 1.4 |
|  | NT (mgN/L) | 8.5 | 8.0 | 9.8 | 11 | 6.7 |
| Primary sludge (2) | TSS (g/L) | 5.54 | 1.50 | 11.4 | 14.5 | 9.33 |
|  | VSS (g/L) | 2.93 | 1.10 | 8.89 | 10.2 | 6.22 |
| Biologic Sludge A (5) | TSS (g/L) | 6.15 | 6.87 | 9.00 | 5.43 | 7.63 |
|  | VSS (g/L) | 4.29 | 5.12 | 7.00 | 4.05 | 5.60 |
| Biologic Sludge B (6) | TSS (g/L) | 6.34 | 5.64 | 6.68 | 5.67 | 6.31 |
|  | VSS (g/L) | 4.29 | 4.52 | 4.33 | 4.12 | 4.59 |
| Thickened Sludge A (7) | TSS (g/L) | 28.2 | 27.3 | 26.1 | 20.8 | NA |
|  | VSS (g/L) | 20.8 | 20.1 | 20.2 | 15.1 | NA |
| Thickened Sludge B (8) | TSS (g/L) | 28.7 | 16.9 | 25.1 | 19.2 | 17.5 |
|  | VSS (g/L) | 20.6 | 19.8 | 19.1 | 13.8 | 10.5 |
| Digested Sludge A (9) | TSS (g/L) | 43.8 | 41.4 | 41.4 | 44.0 | 43.7 |
|  | VSS (g/L) | 23.8 | 21.7 | 22.2 | 23.6 | 23.6 |
| Digested Sludge B (10) | TSS (g/L) | 40.8 | 38.7 | 37.9 | 38.5 | 41.2 |
|  | VSS (g/L) | 23.0 | 21.1 | 20.2 | 21.5 | 22.5 |

Indeed, the influence of rain on sewerage streams is a hurdle to the use of the WWTP influent as an epidemiological indicator. However, the sludge streams tend to have a more steady content of biosolids as it reflects the mass flow of solids and COD entering the WWTP. Particularly, solid concentration in thickeners seems to be much more stable than the characteristics of the primary influent.

### Presence of SARS-CoV-2 genetic material in water and sludge

A total of 15 samples of water and 35 samples of sludge collected from April 6 to April 21 (2020) were tested for the presence of SARS-CoV-2 RNA. All samples were positive for our internal control, bacteriophage MS2, although with variable efficiency (33.3 % ± 15.6). Such a variation has been described before (Petterson et al., 2015; Silva-Sales et al., 2020), probably due to the complexity and variability of sewage samples.

The interpretation of results in Table 3 (complete samples in table A1) was based on considering a positive sample when the cycle threshold took place below cycle 40, for either RNA-dependent polymerase (RdRP) and nucleocapsid (N) SARS-CoV-2 specific genes. Sole detection of gene E, characteristic of pan-Sarbecoviruses prompted for repeating the analysis for confirmation and usually led to a negative result. For most entries in table 3, both specific genes were detected, suggesting that non-specific amplification is unlikely. Yet and being complex samples, sequencing the product of PCR would rule out this risk and confirm the validity of the method for wastewater and sludge. Focusing on the viral loadings, it is important to bear in mind that the most of the cycle thresholds are close to 40, hence close to the limit of detection in the current experimental configuration

**Table 3.** RT-PCR as mean amplification cycles for RNA-dependent polymerase (RdRP) and nucleocapsid (N) SARS-CoV-2 specific genes, a conserved region in of the structural protein envelope for the detection of pan-Sarbecoviruses (E). ND stands for a non-detected gene. Only positive samples are shown in this table (full table in Supplementary Material). The (number) after the sampling spot corresponds to figure 1.

| Sampling spot | Ct E | Ct RdRp | Ct N |
| --- | --- | --- | --- |
| **April 6** |  |  |  |
| Wastewater (1) | ND | ND | 39.60 |
| Wastewater (1) | ND | 37.05 | ND |
| Mixed thickened sludge –A (7) | 35.50 | ND | 38.00 |
| **April 7** |  |  |  |
| Wastewater (1) | ND | 37.57 | 38.84 |
| Primary sludge (2) | 34.34 | 36.02 | 39.87 |
| Sec. treatment sludge – Reactor 1 (5) | ND | 37.14 | 39.35 |
| Mixed thickened sludge –A (7) | 34.66 | ND | 36.15 |
| Mixed thickened sludge –B (8) | 34.48 | ND | 38.28 |
| **April 14** |  |  |  |
| Wastewater (1) | 33.79 | 35.01 | 37.13 |
| Wastewater (1) | 33.83 | 35.46 | 36.39 |
| Primary sludge (2) | ND | ND | 38.38 |
| Mixed thickened sludge –A (7) | 33.79 | ND | 36.19 |
| Mixed thickened sludge –B (8) | 34.73 | 38.08 | 36.62 |
| **April 16** |  |  |  |
| Wastewater (1) | 33.61 | 36.06 | 38.5 |
| Wastewater (1) | 33.64 | 34.7 | 38.08 |
| Primary sludge (2) | 33.41 | 35.92 | 36.62 |
| Mixed thickened sludge –A (7) | 36.09 | 34.77 | 37.21 |
| Mixed thickened sludge –B (8) | 36.13 | ND | 38.21 |
| **April 21** |  |  |  |
| Wastewater (1) | 34.87 | 35.5 | 37.42 |
| Primary sludge (2) | 34.40 | ND | 35.78 |
| Outflow primary settler (3) | 34.09 | 37.68 | 37.31 |
| Mixed thickened sludge –A (7) | 34.66 | ND | 39.34 |
| Mixed thickened sludge –B (8) | 34.62 | 37.19 | 39.03 |
| Mixed thickened sludge –B (8) | 35.18 | 36.53 | 37.43 |

### Quantification of SARS-CoV-2 particles in water and sludge line WWTP

Following the presence of SARS-CoV-2 genetic material along the WWTP allows inferring its fate in the different processes. For that purpose, samples were quantified using commercial standards, as explained in M&M section, leading to the results shown in Table 4.

**Table 4.** Quantification of SARS-CoV-2 genetic material at several WWTP sampling spots. (Pos = positive; Neg = negative; Inh = inhibited). Coloured legend in cp/ml

| OURENSE WWTP |  | 6-Apr | 7-Apr | 14-Apr | 16-Apr | 21-Apr |
| --- | --- | --- | --- | --- | --- | --- |
| Water line | Influent (1) | Pos | Pos | Pos | Pos | Pos |
|  | Outflow Primary (3) | Inh | Neg | Neg | Neg | Pos |
|  | Treated Effluent(4) | Neg | Neg | Neg | Neg | Neg |
| Sludge line | Primary sludge(2) | Neg | Pos | Pos | Pos | Pos |
|  | Biologic Sludge A (5) | Neg | Pos | Neg | Neg | Neg |
|  | Biologic Sludge B(6) | Inh | Neg | Neg | Neg | Neg |
|  | Thickened Sludge A (7) | Pos | Pos | Pos | Pos | Pos |
|  | Thickened Sludge B (8) | Neg | Pos | Pos | Pos | Pos |
|  | Digested Sludge A (9) | Neg | Neg | Neg | Neg | Neg |
|  | Digested Sludge B (10) | Neg | Neg | Neg | Neg | Neg |
| Cases | Total number | 999 | 1001 | 1193 | 1207 | 1301 |
|  | Cases/100 000 | 324.4 | 325.0 | 387.3 | 391.9 | 422.4 |
| Negative<br><7.5<br><10<br><15<br><20<br><40 |  |  |  |  |  |  |

SARS-CoV-2 RNA was systematically detected in the influent to the primary settler (between 7.5 and 15 cp/ml) but not in the secondary treatment effluent, confirming that the effluent is safe for reuse and discharge to water bodies, as other studies have also reported (Randazzo et al., 2020b). Another potential mechanism of transmission of airborne is the production of aerosol in secondary treatment, particularly if aeration is provided by horizontal rotors or surface turbines (Gotkowska-Pƚachta et al., 2013; Sánchez-Monedero et al., 2008). Given the rare occurrence of SARS-CoV-2 RNA in the inflow to the secondary treatment, the potential of dispersion by aerosols created during aeration can be ruled out.

As for the sludge line, it appears that SARS-CoV-2 RNA is mainly retained at the primary settler (10 - 40 cp/ml) and only detected in one occasion in the biological sludge (7.5-10 cp/ml), which suggest that, as hypothesized, the virus particles have a higher affinity for the sludge and therefore, it is mostly diverted to the sludge line. Interestingly, its concentration increases in the thickeners which have a longer retention time, (approximately 24 hours) and a higher solid content.

However, no genetic material is detected in the digested sludge, which is surely related both to the severe temperature undergone during thermal hydrolysis and to the long residence time in the anaerobic digesters. Therefore, the results confirm the safety of the sludge after thermal treatment and anaerobic digestion. However, in smaller WWTPs is only treated by volume reduction methods with no thermal treatment, the safety of sludge disposal remains to be verified.

### Primary and/or thickened sludge as indicators of incidence

It is seen that the concentration of SARS-CoV-2 genetic material is systematically higher in some sludge sampling spots (particularly at primary sludge and thickened sludge) compared to the influent samples (Table 4). This result confirms one of the hypothesis of this work, namely that the affinity of virus particles for biosolids would divert the genetic material of SARS-CoV-2 towards the sludge line. Although the number of samples analysed in this work is limited and replication in other WWTPs is required, this finding suggests that monitoring COVID-19 incidence in the population in the sludge might have a higher sensitivity than in the wastewater. In this WWTP, the primary settler and the sludge thickeners would act in effect as "concentrators" of SARS-CoV-2 genetic material. Furthermore, the retention times in sludge thickeners (~24h) are usually higher than in primary settlers (~1-2 h). This higher retention time results in dampening the potential variations of SARS-CoV-2 particles in wastewater in an even more effective way than taking composite samples. Such a buffering is not helpful when the phenomenon to be monitored has fast dynamics, but in the case of COVID-19 population incidence, the desired monitoring dynamics would be in the order of days, making it a suitable sampling spot.

## Conclusions

The affinity of SARS-CoV-2 by biosolids was seen to govern to a large extent its fate in WWTPs by being associated to sludge streams. As a consequence, SARS-CoV-2 genetic material was not detected in the water effluent, confirming its safety. The combined treatment of thermal hydrolysis and anaerobic digestion also prevented the detection of SARS-CoV-2 in sludge leaving the plant. The primary sludge and mostly the thickened sludge showed higher and steadier concentrations, which suggests that COVID-19 incidence could be monitored preferably in the sludge line rather than, or in addition to, the raw wastewater. Longer residence times and higher solid concentrations in sludge thickeners would make it a more robust sampling spot, which merits being further investigated.

## Data Availability

All the data are available in the manuscript body or in supplementary material provided with the manuscript

## Acknowledgements

Sabela Balboa, Miguel Mauricio-Iglesias and Juan M. Lema belong to the CRETUS Strategic Partnership (ED431E 2018/01) and to the Galician Competitive Research Group (ED431C2017/029). All these programs are co-funded by ERDF (EU).

## Annex 1. Supplementary Material

The supplementary material contains the results of the RT-PCR and physicochemical characteristics of the water and sludge for all the sampling points and days.

**Table A1.** Full collection of samples, showing results of RT-PCR as mean amplification cycles for RNA-dependent polymerase (RdRP) and nucleocapsid (N) SARS-CoV-2 specific genes, a conserved region in of the structural protein envelope for the detection of pan-Sarbecoviruses (E). The (number) after the sampling spot corresponds to figure 1. ND stands for a non-detected gene.

| Sampling spot | Ct E | Ct RdRp | Ct N | Interpretation |
| --- | --- | --- | --- | --- |
| **April 6** |  |  |  |  |
| Wastewater (1) | ND | ND | 39.6 | Positive |
| Wastewater (1) | ND | 37.05 | ND | Positive |
| Primary sludge (2) | ND | ND | ND | Negative |
| Primary sludge (2) | ND | ND | ND | Negative |
| Outflow primary settler (3) |  |  |  | Inhibited |
| Outflow sec. treatment (4) | ND | ND | ND | Negative |
| Outflow sec. treatment (4) | ND | ND | ND | Negative |
| Sec. treatment sludge – Reactor 1 (5) | ND | ND | ND | Negative |
| Sec. treatment sludge – Reactor 2 (6) |  |  |  | Inhibited |
| Mixed thickened sludge –A (7) | 35.5 | X | 38.0 | Positive |
| Mixed thickened sludge –B (8) | ND | ND | ND | Negative |
| Digested sludge – A (9) | ND | ND | ND | Negative |
| Digested sludge – B (10) | ND | ND | ND | Negative |
| **April 7** |  |  |  |  |
| Wastewater (1) | ND | ND | ND | Negative |
| Wastewater (1) | ND | 37.57 | 38.84 | Positive |
| Primary sludge (2) | 34.34 | 36.02 | 39.87 | Positive |
| Outflow primary settler (3) | ND | ND | ND | Negative |
| Outflow primary settler (3) | ND | ND | ND | Negative |
| Outflow sec. treatment (4) | ND | ND | ND | Negative |
| Outflow sec. treatment (4) | ND | ND | ND | Negative |
| Sec. treatment sludge – Reactor 1 (5) | ND | 37.14 | 39.35 | Positive |
| Sec. treatment sludge – Reactor 2 (6) | ND | ND | ND | Negative |
| Mixed thickened sludge –A (7) | 34.66 | ND | 36.15 | Positive |
| Mixed thickened sludge –B (8) | 34.48 | ND | 38.28 | Positive |
| Digested sludge – A (9) | ND | ND | ND | Negative |
| Digested sludge – B (10) | ND | ND | ND | Negative |

| **April 14** |  |  |  |  |
| --- | --- | --- | --- | --- |
| Wastewater (1) | 33.79 | 35.01 | 37.13 | Positive |
| Wastewater (1) | 33.83 | 35.46 | 36.39 | Positive |
| Primary sludge (2) | ND | ND | 38.38 | Positive |
| Outflow primary settler (3) | ND | ND | ND | Negative |
| Outflow primary settler (3) | ND | ND | ND | Negative |
| Outflow sec. treatment (4) | ND | ND | ND | Negative |
| Outflow sec. treatment (4) | ND | ND | ND | Negative |
| Sec. treatment sludge - Reactor 1 (5) | ND | ND | ND | Negative |
| Sec. treatment sludge - Reactor 2 (6) | ND | ND | ND | Negative |
| Mixed thickened sludge -A (7) | 33.79 | ND | 36.19 | Positive |
| Mixed thickened sludge -B (8) | 34.73 | 38.08 | 36.62 | Positive |
| Digested sludge - A (9) | ND | ND | ND | Negative |
| Digested sludge - B (10) | ND | ND | ND | Negative |
| **April 16** |  |  |  |  |
| Wastewater (1) | 33.61 | 36.06 | 38.5 | Positive |
| Wastewater (1) | 33.64 | 34.7 | 38.08 | Positive |
| Primary sludge (2) | 33.41 | 35.92 | 36.62 | Positive |
| Outflow primary settler (3) | ND | ND | ND | Negative |
| Outflow primary settler (3) | ND | ND | ND | Negative |
| Outflow sec. treatment (4) | ND | ND | ND | Negative |
| Outflow sec. treatment (4) | ND | ND | ND | Negative |
| Sec. treatment sludge – Reactor 1 (5) | ND | ND | ND | Negative |
| Sec. treatment sludge – Reactor 2 (6) | ND | ND | ND | Negative |
| Mixed thickened sludge –A (7) | 36.09 | 34.77 | 37.21 | Positive |
| Mixed thickened sludge –B (8) | 36.13 | ND | 38.21 | Positive |
| Digested sludge – A (9) | ND | ND | ND | Negative |
| Digested sludge – B (10) | ND | ND | ND | Negative |
| **April 21** |  |  |  |  |
| Wastewater (1) | 34.87 | 35.5 | 37.42 | Positive |
| Primary sludge (2) | 34.4 | ND | 35.78 | Positive |
| Outflow primary settler (3) | 34.09 | 37.68 | 37.31 | Positive |
| Sec. treatment sludge - Reactor 1 (5) | ND | ND | ND | Negative |
| Sec. treatment sludge - Reactor 2 (6) | ND | ND | ND | Negative |
| Mixed thickened sludge -A (7) | 34.66 | ND | 39.34 | Positive |
| Mixed thickened sludge -A (7) | ND | ND | ND | Negative |
| Mixed thickened sludge -B (8) | 34.62 | 37.19 | 39.03 | Positive |
| Mixed thickened sludge -B (8) | 35.18 | 36.53 | 37.43 | Positive |
| Digested sludge - A (9) | ND | ND | ND | Negative |
| Digested sludge - B (10) | ND | ND | ND | Negative |

**Table A2.** Samples physicochemical characterisation (April 6)

|  | Wastewater<br>(1) | Outflow<br>primary<br>settler<br>(3) | Outflow<br>sec.<br>treatment<br>(4) | Primary<br>sludge<br>(2) | Sec.<br>treatment<br>sludge -<br>Reactor 1<br>(5) | Sec.<br>treatment<br>sludge -<br>Reactor 2<br>(6) | Mixed<br>thickened<br>sludge -A<br>(7) | Mixed<br>thickened<br>sludge -A<br>(8) | Digested<br>sludge -<br>A (9) | Digested<br>sludge - B<br>(10) |
| --- | --- | --- | --- | --- | --- | --- | --- | --- | --- | --- |
| **pH** | 7.01 | 7.46 | 6.76 | 6.04 | 6.33 | 6.38 | 6.48 | 6.44 | 7.83 | 7.51 |
| **Conductivity**<br>**(µS/cm)** | 440 | 508 | 294 |  |  |  |  |  | 13 170 | 10 910 |
| **TSS (mg/L)** | 992.00 | 51.00 | 2.50 | 5 546 | 6 148 | 6 434 | 28 292 | 28 668 | 43 848 | 40 752 |
| **VSS (mg/L)** |  |  |  | 2 934 | 4 286 | 4 292 | 20 824 | 20 636 | 23 804 | 22 956 |
| **COD (mg/L)** | 1211 | 101 | 17 |  |  |  |  |  |  |  |
| **NH4 - N (mg/L)** | 26.70 | 25.40 | 1.60 |  |  |  |  |  |  |  |
| **NH4 (mg/L)** | 34.23 | 32.56 | 2.05 |  |  |  |  |  |  |  |
| **N - Total (mg/L)** | 56.00 | 35.00 | 8.50 |  |  |  |  |  |  |  |
| **P - Total (mg/L)** | 13.00 | 3.10 | 0.31 |  |  |  |  |  |  |  |

**Table A3.** Samples physicochemical characterisation (April 7)

|  | Wastewater<br>(1) | Outflow<br>primary<br>settler<br>(3) | Outflow<br>sec.<br>treatment<br>(4) | Primary<br>sludge<br>(2) | Sec.<br>treatment<br>sludge –<br>Reactor 1<br>(5) | Sec.<br>treatment<br>sludge –<br>Reactor 2<br>(6) | Mixed<br>thickened<br>sludge –A<br>(7) | Mixed<br>thickened<br>sludge –A<br>(8) | Digested<br>sludge – A<br>(9) | Digested<br>sludge – B<br>(10) |
| --- | --- | --- | --- | --- | --- | --- | --- | --- | --- | --- |
| **pH** | 7.01 | 7.46 | 6.76 | 6.10 | 6.44 | 6.41 | 6.50 | 6.47 | 7.81 | 7.67 |
| **Conductivity**<br>**(µS/cm)** | 362 | 397 | 428 |  |  |  |  |  |  |  |
| **TSS (mg/L)** | 105.00 | 75.00 | 6.00 | 1 502 | 6 870 | 6 642 | 27 300 | 26 872 | 41 396 | 38 672 |
| **VSS (mg/L)** |  |  |  | 1 102 | 5 176 | 4 524 | 20 100 | 19 772 | 21 740 | 21 132 |
| **COD (mg/L)** | 64 | 53 | 13 |  |  |  |  |  |  |  |
| **NH4 - N (mg/L)** | 13.80 | 18.60 | 0.90 |  |  |  |  |  |  |  |
| **N - Total (mg/L)** | 21.00 | 26.00 | 8.00 |  |  |  |  |  |  |  |
| **P - Total (mg/L)** | 2.10 | 2.00 | 0.52 |  |  |  |  |  |  |  |

**Table A4.** Samples physicochemical characterisation (April 14)

|  | Wastewater<br>(1) | Outflow<br>primary<br>settler<br>(3) | Outflow<br>sec.<br>treatment<br>(4) | Primary<br>sludge<br>(2) | Sec.<br>treatment<br>sludge -<br>Reactor 1<br>(5) | Sec.<br>treatment<br>sludge -<br>Reactor 2<br>(6) | Mixed<br>thickened<br>sludge -A<br>(7) | Mixed<br>thickened<br>sludge -A<br>(8) | Digested<br>sludge -<br>A (9) | Digested<br>sludge - B<br>(10) |
| --- | --- | --- | --- | --- | --- | --- | --- | --- | --- | --- |
| **pH** | 7.23 | 7.38 | 7.13 | 6.75 | 6.30 | 6.57 | 6.66 | 6.69 | 7.72 | 7.48 |
| **Conductivity**<br>**(µS/cm)** | 539 | 527 | 363 |  |  |  |  |  |  |  |
| **TSS (mg/L)** | 284.00 | 128.00 | 8.60 | 11 396 | 9 002 | 6 682 | 26 100 | 25 056 | 41 448 | 37 916 |
| **VSS (mg/L)** |  |  |  | 8 888 | 7 001 | 4 336 | 20 188 | 19 089 | 22 180 | 20 240 |
| **COD (mg/L)** | 613 | 212 | 23 |  |  |  |  |  |  |  |
| **NH4 - N (mg/L)** | 35.50 | 31.90 | 1.30 |  |  |  |  |  |  |  |
| **N - Total (mg/L)** | 52.00 | 41.00 | 9.80 |  |  |  |  |  |  |  |
| **P - Total (mg/L)** | 6.00 | 4.40 | 0.58 |  |  |  |  |  |  |  |

**Table A5.**
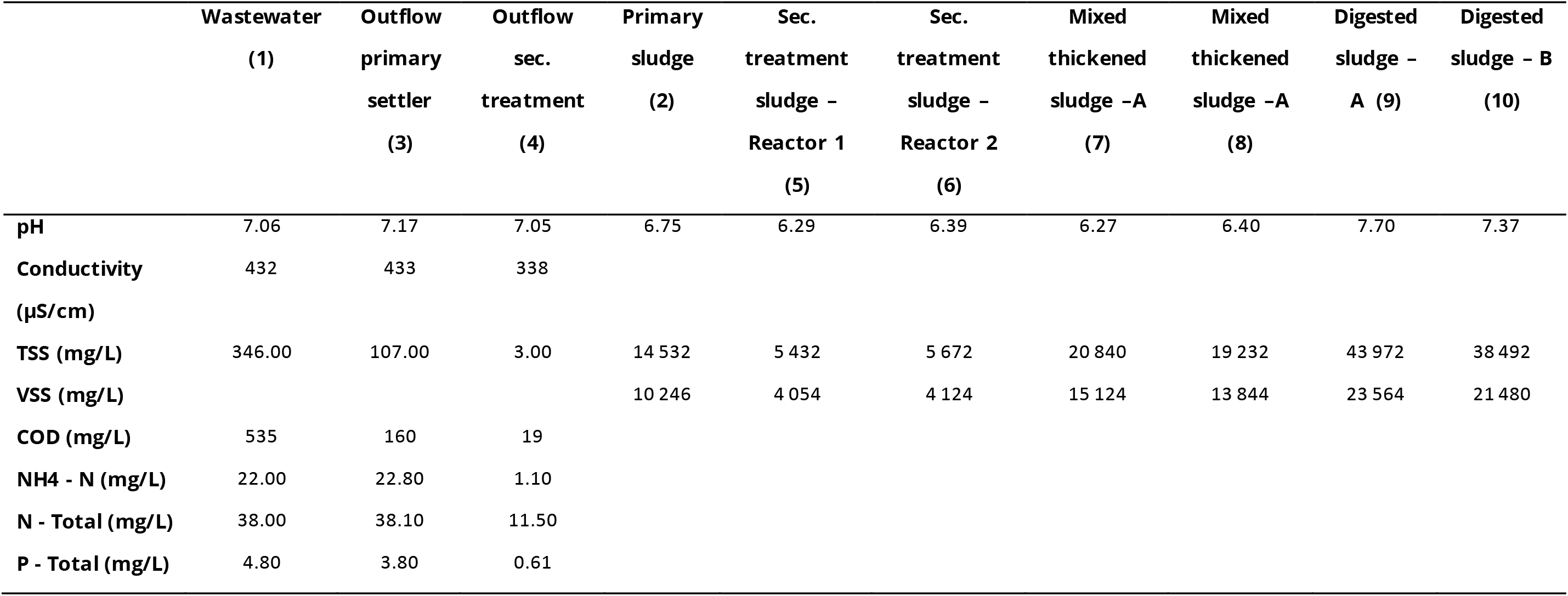
Samples physicochemical characterisation (April 16)

|  | Wastewater<br>(1) | Outflow<br>primary<br>settler<br>(3) | Outflow<br>sec.<br>treatment<br>(4) | Primary<br>sludge<br>(2) | Sec.<br>treatment<br>sludge -<br>Reactor 1<br>(5) | Sec.<br>treatment<br>sludge -<br>Reactor 2<br>(6) | Mixed<br>thickened<br>sludge -A<br>(7) | Mixed<br>thickened<br>sludge -A<br>(8) | Digested<br>sludge -<br>A (9) | Digested<br>sludge - B<br>(10) |
| --- | --- | --- | --- | --- | --- | --- | --- | --- | --- | --- |
| **pH** | 7.06 | 7.17 | 7.05 | 6.75 | 6.29 | 6.39 | 6.27 | 6.40 | 7.70 | 7.37 |
| **Conductivity**<br>**(µS/cm)** | 432 | 433 | 338 |  |  |  |  |  |  |  |
| **TSS (mg/L)** | 346.00 | 107.00 | 3.00 | 14 532 | 5 432 | 5 672 | 20 840 | 19 232 | 43 972 | 38 492 |
| **VSS (mg/L)** |  |  |  | 10 246 | 4 054 | 4 124 | 15 124 | 13 844 | 23 564 | 21 480 |
| **COD (mg/L)** | 535 | 160 | 19 |  |  |  |  |  |  |  |
| **NH4 - N (mg/L)** | 22.00 | 22.80 | 1.10 |  |  |  |  |  |  |  |
| **N - Total (mg/L)** | 38.00 | 38.10 | 11.50 |  |  |  |  |  |  |  |
| **P - Total (mg/L)** | 4.80 | 3.80 | 0.61 |  |  |  |  |  |  |  |

**Table A6.**
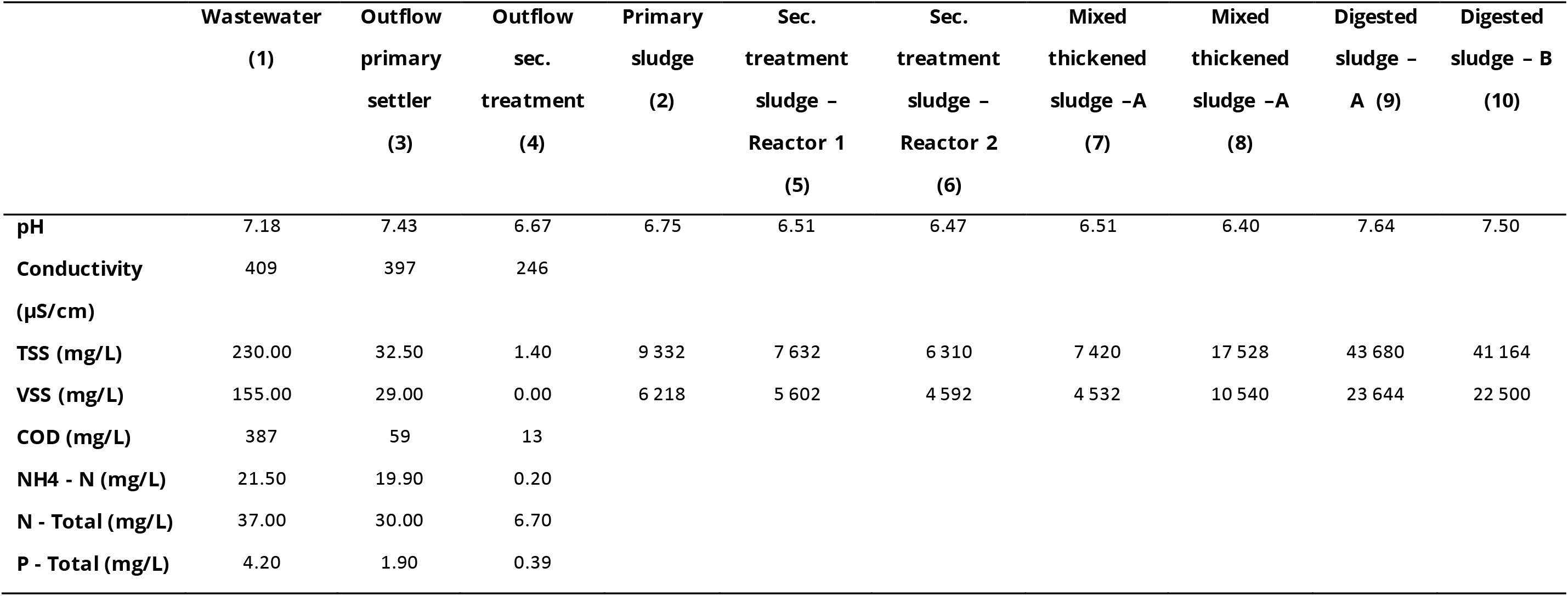
Samples physicochemical characterisation (April 21)

|  | Wastewater<br>(1) | Outflow<br>primary<br>settler<br>(3) | Outflow<br>sec.<br>treatment<br>(4) | Primary<br>sludge<br>(2) | Sec.<br>treatment<br>sludge -<br>Reactor 1<br>(5) | Sec.<br>treatment<br>sludge -<br>Reactor 2<br>(6) | Mixed<br>thickened<br>sludge -A<br>(7) | Mixed<br>thickened<br>sludge -A<br>(8) | Digested<br>sludge -<br>A (9) | Digested<br>sludge - B<br>(10) |
| --- | --- | --- | --- | --- | --- | --- | --- | --- | --- | --- |
| **pH** | 7.18 | 7.43 | 6.67 | 6.75 | 6.51 | 6.47 | 6.51 | 6.40 | 7.64 | 7.50 |
| **Conductivity**<br>**(µS/cm)** | 409 | 397 | 246 |  |  |  |  |  |  |  |
| **TSS (mg/L)** | 230.00 | 32.50 | 1.40 | 9 332 | 7 632 | 6 310 | 7 420 | 17 528 | 43 680 | 41 164 |
| **VSS (mg/L)** | 155.00 | 29.00 | 0.00 | 6 218 | 5 602 | 4 592 | 4 532 | 10 540 | 23 644 | 22 500 |
| **COD (mg/L)** | 387 | 59 | 13 |  |  |  |  |  |  |  |
| **NH4 - N (mg/L)** | 21.50 | 19.90 | 0.20 |  |  |  |  |  |  |  |
| **N - Total (mg/L)** | 37.00 | 30.00 | 6.70 |  |  |  |  |  |  |  |
| **P - Total (mg/L)** | 4.20 | 1.90 | 0.39 |  |  |  |  |  |  |  |

## References

DIN, 2011. 38405–9:2011-09 German standard methods for examination of water, waste water and sludge - Anions (group D) - Part 9: Spectrometric determination of nitrate (D 9).

Ehlers, M.M., Grabow, W.O.K., Pavlov, D.N., 2005. Detection of enteroviruses in untreated and treated drinking water supplies in South Africa. Water Res. 39, 2253–2258. https://doi.org/10.1016/j.watres.2005.04.014

Gotkowska-Płachta, A., Filipkowska, Z., Korzeniewska, E., Janczukowicz, W., Dixon, B., Goƚas, I., Szwalgin, D., 2013. Airborne Microorganisms Emitted from Wastewater Treatment Plant Treating Domestic Wastewater and Meat Processing Industry Wastes. CLEAN - Soil, Air, Water 41, 429–436. https://doi.org/10.1002/clen.201100466

Hjelmsø, M.H., Hellmér, M., Fernandez-Cassi, X., Timoneda, N., Lukjancenko, O., Seidel, M., Elsässer, D., Aarestrup, F.M., Löfström, C., Bofill-Mas, S., Abril, J.F., Girones, R., Schultz, A.C., 2017. Evaluation of methods for the concentration and extraction of viruses from sewage in the context of metagenomic sequencing. PLoS One 12, 1–17. https://doi.org/10.1371/journal.pone.0170199

Holshue, M.L., DeBolt, C., Lindquist, S., Lofy, K.H., Wiesman, J., Bruce, H., Spitters, C., Ericson, K., Wilkerson, S., Tural, A., Diaz, G., Cohn, A., Fox, L.A., Patel, A., Gerber, S.I., Kim, L., Tong, S., Lu, X., Lindstrom, S., Pallansch, M.A., Weldon, W.C., Biggs, H.M., Uyeki, T.M., Pillai, S.K., 2020. First case of 2019 novel coronavirus in the United States. N. Engl. J. Med. 382, 929–936. https://doi.org/10.1056/NEJMoa2001191

International Organization for Standardization, 1997. ISO 11905–1:1997 Water quality — Determination of nitrogen — Part 1: Method using oxidative digestion with peroxodisulfate.

Medema, G., Heijnen, L., Elsinga, G., Italiaander, R., Medema, G., 2020. Presence of SARS-Coronavirus-2 in sewage. Methods Sewage samples. medRxiv. https://doi.org/10.1101/2020.03.29.20045880

Petterson, S., Grøndahl-Rosado, R., Nilsen, V., Myrmel, M., Robertson, L.J., 2015. Variability in the recovery of a virus concentration procedure in water: Implications for QMRA. Water Res. 87, 79–86. https://doi.org/10.1016/j.watres.2015.09.006

Prado, T., Gaspar, A.M.C., Miagostovich, M.P., 2014. Detection of enteric viruses in activated sludge by feasible concentration methods. Braz. J. Microbiol. 45, 343–349. https://doi.org/10.1590/s1517-83822014000100049

Randazzo, W., Cuevas-Ferrando, E., Sanjuan, R., Domingo-Calap, P., Sanchez, G., 2020a. Metropolitan Wastewater Analysis for COVID-19 Epidemiological Surveillance. medRxiv 2020.04.23.20076679. https://doi.org/10.1101/2020.04.23.20076679

Randazzo, W., Truchado, P., Ferrando, E.C., Simon, P., Allende, A., Sanchez, G., 2020b. SARS-CoV-2 RNA titers in wastewater anticipated COVID-19 occurrence in a low prevalence area. Water Res. https://doi.org/10.1016/j.watres.2020.115942

Rosa, G. La Iaconelli, M., Mancini, P., Ferraro, G.B., Veneri, C., Bonadonna, L., Lucentini, L., 2020. First Detection of Sars-Cov-2 in Untreated Wastewaters in Italy. medRxiv 2020.04.25.20079830. https://doi.org/10.1101/2020.04.25.20079830

Sánchez-Monedero, M.A., Aguilar, M.I., Fenoll, R., Roig, A., 2008. Effect of the aeration system on the levels of airborne microorganisms generated at wastewater treatment plants. Water Res. 42, 3739–3744. https://doi.org/10.1016/j.watres.2008.06.028

Sassi, H.P., Tuttle, K.D., Betancourt, W.Q., Kitajima, M., Gerba, C.P., 2018. Persistence of Viruses by qPCR Downstream of Three Effluent-Dominated Rivers in the Western United States. Food Environ. Virol. 10, 297–304. https://doi.org/10.1007/s12560-018-9343-7

Silva-Sales, M., Martínez-Puchol, S., Gonzales-Gustavson, E., Hundesa, A., Gironès, R., 2020. High Prevalence of Rotavirus A in Raw Sewage Samples from Northeast Spain. Viruses 12, 318. https://doi.org/10.3390/v12030318

Symonds, E.M., Sinigalliano, C., Gidley, M., Ahmed, W., McQuaig-Ulrich, S.M., Breitbart, M., 2016. Faecal pollution along the southeastern coast of Florida and insight into the use of pepper mild mottle virus as an indicator. J. Appl. Microbiol. 121, 1469–1481. https://doi.org/10.1111/jam.13252

Taboada-Santos, A., Rivadulla, E., Paredes, L., Carballa, M., Romalde, J., Lema, J.M., 2020. Comprehensive comparison of chemically enhanced primary treatment and high-rate activated sludge in novel wastewater treatment plant configurations. Water Res. 169, 115258. https://doi.org/10.1016/j.watres.2019.115258

Wellings, F.M., Lewis, A.L., Mountain, C.W., 1976. Demonstration of solids-associated virus in wastewater and sludge. Appl. Environ. Microbiol. 31, 354–358.

Woelfel, R., Corman, V.M., Guggemos, W., Seilmaier, M., Zange, S., Mueller, M.A., Niemeyer, D., Vollmar, P., Rothe, C., Hoelscher, M., Bleicker, T., Bruenink, S., Schneider, J., Ehmann, R., Zwirglmaier, K., Drosten, C., Wendtner, C., 2020. Virological assessment of hospitalized cases of coronavirus disease 2019. medRxiv 2020.03.05.20030502. https://doi.org/10.1101/2020.03.05.20030502

Wu, F., Xiao, A., Zhang, J., Gu, X., Lee, W.L., Kauffman, K., Hanage, W., Matus, M., Ghaeli, N., Endo, N., Duvallet, C., Moniz, K., Erickson, T., Chai, P., Thompson, J., Alm, E., 2020. SARS-CoV-2 titers in wastewater are higher than expected from clinically confirmed cases. medRxiv 2020.04.05.20051540. https://doi.org/10.1101/2020.04.05.20051540

Wurtzer, S., Marechal, V., Mouchel, J.-M., Moulin, L., 2020. Time course quantitative detection of SARS-CoV-2 in Parisian wastewaters correlates with COVID-19 confirmed cases. medRxiv 2020.04.12.20062679. https://doi.org/10.1101/2020.04.12.20062679

Ye, Y., Ellenberg, R.M., Graham, K.E., Wigginton, K.R., 2016. Survivability, Partitioning, and Recovery of Enveloped Viruses in Untreated Municipal Wastewater. Environ. Sci. Technol. 50, 5077–5085. https://doi.org/10.1021/acs.est.6b00876

Yu, I.T.S., Li, Y., Wong, T.W., Tam, W., Chan, A.T., Lee, J.H.W., Leung, D.Y.C., Ho, T., 2004. Evidence of Airborne Transmission of the Severe Acute Respiratory Syndrome Virus. N. Engl. J. Med. https://doi.org/10.1056/NEJMoa032867

